# Direct, Indirect and Mixed Methods of Health Education By Nurse and Its Impact on Type 2 Diabetes Patients: A Literature review

**DOI:** 10.1101/2021.12.22.21268287

**Authors:** Emilia Erningwati Akoit, Moses Glorino Rumambo Pandin

## Abstract

**Background:** Diabetes Mellitus (DM) is one of the chronic non-communicable diseases that has currently been very common, in particular Diabetes Mellitus type 2 that threatens public health. It has been included in the category of the six biggest causes of worldwide death, but self-control of treatment and obedience to self-care is still low. One of the influencing factors is related to the lack of knowledge. Providing ongoing health education is one of the solutions or efforts to strengthen knowledge in type 2 diabetes. **The aim** was to identify the various health education methods currently used by nurses and their impacts on type 2 DM. The method used is a literature review. The literature was searched on data based on Scopus, Web of Science, SAGE, CINAHL with the keywords “methods” or “interventions”, “education”, “health”, “nursing”, and “type 2 diabetes melitus. Fifteen (15) pieces of literature were considered to meet the criteria inclusion.

**Results:** Three (3) types of health education methods used by nurses were identified: 1). Direct health education refers to providing education by nurses to patients through training, coaching, interviews, Focus Group Discussion (FGD) and home visits; 2). Indirect Health Education - using mobile phones; 3). The mixed of direct health education and the use of mobile phone-based applications is carried out with the application of mobile health technology and nurse health coaching. The impact of providing health education by nurses to type 2 diabetic patients: increasing of knowledge, behavioral change on preventing diabetes complications, increasing self-efficacy, increasing self-care activities (diet management, physical activity, monitoring blood sugar levels, and foot care).

**Conclusion:** various methods of health education carried out by nurses currently have a positive impact on improving and increasing self-care management and efforts to prevent complications in type 2 diabetes.

## Introduction

Diabetes Mellitus (DM) is one of the chronic diseases that has currently occured and threatened public health. It is characterized by increased blood sugar levels or hyperglycemia due to impaired insulin secretion, insulin action or combination of both. Daibetes, if not handled or controlled properly will have an impact on the risk of various complications - that often occur include disorders of the cardiovascular system, namely hypertension and heart attacks (Lumu *et al*., 2021). Other complications include stroke, kidney failure, blindness, nerve damage or neuropathy that leads to leg or lower limb amputation and reduction of the patient’s life expectancy (Degefa *et al*., 2020).

The prevalence of diabetes, particularly type 2, has continued to increase in the last 3 decades along with lifestyle changes. It is estimated that around 415 million people in the world suffer diabetes (Jiang *et al*., 2021). The International Diabetes Federation in 2019 also predicted that in 2030 the number of people with diabetes will grow to be 578 million and it will increase to 700 million in 2045. In 2018, World Health Organization (WHO) determined diabetes as one of four non-communicable diseases were prioritized for treatment. In Indonesia, the incidence of diabetes continues to increase. The Basic Health Research Report (Riskesdas) in 2018, which is one of the national-scale researches, found that the prevalence of diabetes based on the results of blood sugar examinations showed an increase from 6.9% in 2013 to 8.5% in 2018.

Diabetes is included in the category of the six biggest causes of the worldwide death, but control over treatment and obedience to self-care is still low (Ahrary *et al*., 2020). Several factors that influence compliance were identified. They are lack of knowledge, personal and cultural beliefs, communication and access to health and education services (Goff *et al*., 2021). It was further explained that the level of obedience to diabetical treatment was low at around 30-89.7% (Trevisan *et al*., 2020). These situations contribute an impact on uncontrolled blood glucose levels and the risk of increasing the occurrence of various complications. Therefore, various approaches or management need to be made to increase knowledge, awareness, understanding and patient compliance to diabetic care, including provision of health education.

Health education is an approach or effort to provide information to increase one’s knowledge and understanding in terms of improving health, preventing disease and managing a disease. Diabetic patients need to be educated in order to increase their knowledge, and change their attitudes and behavior in the management of diabetes, particularly for long-term diabetic care. (Moyeda-Carabaza *et al*., 2020) conducted a study on the effect of health education interventions on diabetes and the factors influence it. It demonstrated that health education has a positive impact on the attitudes and behavior of the patients with diabetes type 2 in its management.

Nurse, as one of the health workers who can act as an educator. Nurse can provide health education to increase knowledge, attitudes and behavior of the diabetic patients in self-care (Sousa *et al*., 2020). Effective health education can increase self-confidence, develop new mindsets and increase compliance of the diabetic patients in performing self-care (Sousa *et al*., 2020). Diabetes in terms of diet regulation, physical exercise, medication management, lower limb care and independent monitoring of blood glucose levels (Lie *et al*., 2019).

Methods of health education used by nurses should be varied and innovative that encourage the patient’s improvement to receive the health education that enable diabetic patients to carry out self-care activities independntly. Thus, the quality of nursing services delivered can be further improved.

## Method

The design of this study is a literature review. The literature used in this study consists of research articles related to the methods of health education used by nurse and their impact on type 2 diabetic patients. The articles have been published in reputable international journals. The literature was searched in the data based on Scopus, Web of Science, SAGE, CINAHL with the keywords “method” or “intervention” and “education” and “health” and “nursing” and “type 2 diabetes. In addition, descriptive analysis in the form of narrative is also added.

**Table 1.1.** Inclusion and exclusion criteria with PICOS.

| Criteria | Inclusion | Exclusion |
| --- | --- | --- |
| Population | Type 2 diabetic patients | Type 1 diabetes and the other type of diabetes |
| Intervention | Health education by nurse | Health education by other health professionals or no intervention. |
| Comparisons | There are no comparison | There are no criteria exclusion |
| Outcome | Problem that occur in the type 2 diabetic patients are resolved, for example controlled blood glucose level | Not resolved problem experienced by type 2 diabetic patients |
| Study Type | Randomized Controlled Trial, A quasi experimental, cross sectional, Controlled Clinical Trial, action research, mix method. | Systematic or literature review, qualitative study |
| Publication type | Peer reviewed original studies | Non peer reviewed studies |
| Publication years | 2019-2021 | Pre 2019 |
| Language | English | Other than English |

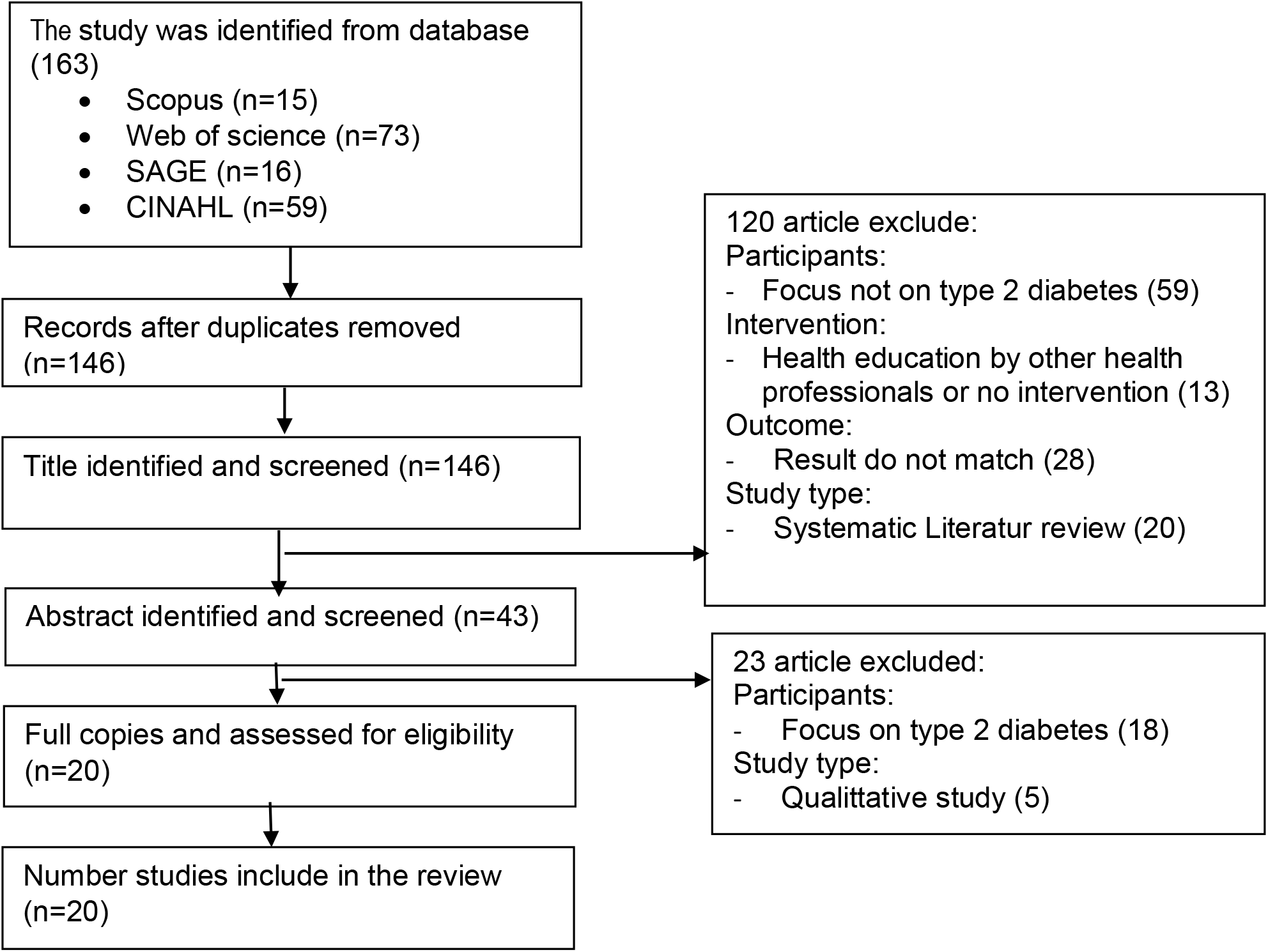

## Result

**Table 1.2.** Distribution of health education direct methods currently used by nurses and their impact on The type 2 diabetes patient

| Researchers, Year | Types of health education direct methods | Impact on the type 2 diabetic patients |
| --- | --- | --- |
| Moyeda-carabaza *et al.*, 2020 | Training session, home visit. | Increased knowledge, positive behavior on the prevention of diabetic complications, increased self-efficacy, decreased excessive intake of carbohydrates and fats. |
| Sousa *et al.*, 2019 | Focus group discussion, interview. | Increased knowledge, increased self-care behavior (diet management, physical activity, monitoring blood sugar levels, lower limb/foot care), increased self-empowerment. |

| **Researchers, Year** | **Types of health education direct methods** | **Impact on the type 2 diabetic patients** |
| --- | --- | --- |
| (Hadziabdic *et al.*, 2020) | Focus Group Discussion (FGD) | Increased knowledge about diabetes, increased self-care behavior and improved health. |
| (Guo *et al.*, 2019) | Educational methods with nurses as health team leaders (use of electronic information systems), distribution of leaflets on diabetic care, support provided in self-care management. | Controlled HbA1c levels, increased knowledge about diabetes and awareness of self-care implementation. |
| (Baptista *et al.*, 2019) | Training, face to face, home visit | Increased knowledge in terms of regulating post prandial blood glucose levels, peripheral blood glucose, diet and medication management, drug administration management for the prevention of hypoglycemia. |
| Ahrary *et al.*, 2020 | Organized hands-on learning | Increased self-care activities, knowledge, attitudes and skills in diabetic management, fasting blood sugar levels and controlled HbA1c. |
| Lumu *et al.*, 2021 | Coaching and lifestyle modification | Decreased systolic blood pressure in type 2 diabetic patients, decreased risk of cardiovascular disease. |
| (Martín-Payo *et al.*, 2021a) | Health education with written guidance and home visits | Increased obedience to recommended diet, activities, decreased HbA1c levels and body mass index. |

**Table 1.3.** Distribution of health education indirect methods currently used by nurses and their impact on The type 2 diabetes patient

| **Researchers, Year** | **Types of health education indirect methods** | **Impact on the type 2 diabetic patients** |
| --- | --- | --- |
| (Asante *et al.*, 2020) | Mobile phone call intervention | Controlled HbA1c levels, increased compliance in self-care (diet, activity/exercise and lower limb/ foot care). |
| (Dincer and Bahçecik, 2021) | Mobile phone application | Increased knowledge, self-efficacy and lower limb/ foot care behavior. |

**Table 1.4.**
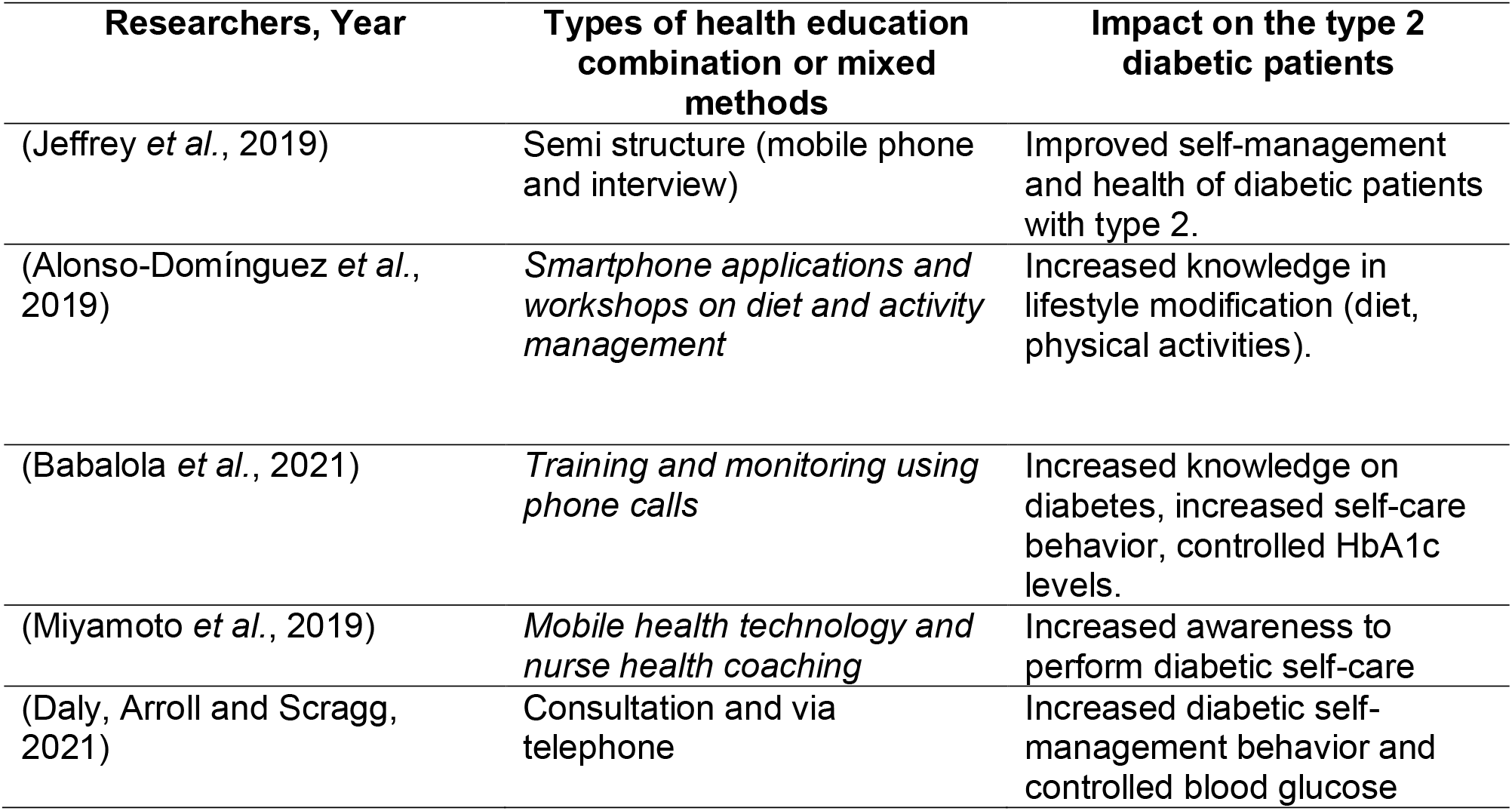
Distribution of health education combination methods currently used by nurses and their impact on type 2 diabetes patient

| **Researchers, Year** | **Types of health education combination or mixed methods** | **Impact on the type 2 diabetic patients** |
| --- | --- | --- |
| (Jeffrey *et al.*, 2019) | Semi structure (mobile phone and interview) | Improved self-management and health of diabetic patients with type 2. |
| (Alonso-Domínguez *et al.*, 2019) | *Smartphone applications and workshops on diet and activity management* | Increased knowledge in lifestyle modification (diet, physical activities). |
| (Babalola *et al.*, 2021) | *Training and monitoring using phone calls* | Increased knowledge on diabetes, increased self-care behavior, controlled HbA1c levels. |
| (Miyamoto *et al.*, 2019) | *Mobile health technology and nurse health coaching* | Increased awareness to perform diabetic self-care |
| (Daly, Arroll and Scragg, 2021) | Consultation and via telephone | Increased diabetic self-management behavior and controlled blood glucose |

## Discussion

Health education is an important approach for nurses to increase knowledge, attitudes and behavior of patients in carrying out care and treatment. In line with the approach, health education conducted by nurses for the type 2 diabetic patients that aims to increase patient knowledge on diabetic management and increasing compliance to self-care activities, thus adequate glycemic control can be achieved. From the study conducted by Trevisan *et al*., 2019 show that health education provided by nurses can increase obidience to therapeutic regimens, which will have a positive effect on blood glucose levels.

### Direct methods and its impact on type 2 diabetic patients

In table 1.1, several direct methods are found to be used by nurses in providing health education to the diabetic patients with type 2 in order to increase knowledge and self-care behavior in reducing the risk of diabetic various complications. Various health education direct methods given to type 2 diabetic patients are a form of nurse innovations to improve the patient’s ability to manage diabetes to achiev optimal glycemic control.

Direct health education can be done through training programs (Martín-Payo *et al*., 2021b); Baptista *et al*., 2019), group discussions - Focus Groups Discussion/FGD (Hadziabdic *et al*., 2020; Sousa *et al*., 2019). Different forms of direct education given to patients is able to increase knowledge, self-care behavior (diet management, physical activity, monitoring blood sugar levels, lower limb/foot care) and self-empowerment. The educational method – group discussion, in particular FGDs consisting of 4-5 people in one group aims to share problems experienced by patients, including the patient’s beliefs in self-care. In FGDs, nurses will be present to provide feedback regarding the things experienced by the patient and supporting the patients with solutions. In addition, health education with written guidelines and home visits can increase adherence to the recommended diet, physical activities and able to reduce HbA1c levels and body mass index (Martín-Payo *et al*., 2021a). The written guidelines contain information on diet and exercise settings for the diabetic management. With written guidelines, patients are easier to follow the nurse’s directions as to increasing the success of the intervention. Furthermore, as explained by Ahrary *et al*., 2020, an organized direct learning will increase self-care activities, knowledge, attitudes and skills in diabetic management, controlled fasting blood sugar levels and HbA1c levels.

The provision of health education by nurses is expected to have a positive impact on the type 2 diabetic patients. Several results from studies demonstrated that providing direct health education enable diabetic patients to accept the material more easily, increasing the patient’s motivation and confidence in practicing self-care activities. Nurse needs to continue to improve health education to patients as a form of playing active role as an educator. Nurse can teach patients about diabetes and its management, including self-care activities and actions to prevent its complications. Health education provided by nurse can increase and strengthen knowledge on diabetes and patient’s awareness on the treatment of the disease (Guo *et al*., 2019).

Nurse has an important role in increasing patient compliance to the management or treatment of any diseases experienced by patients. Supports to improve compliance can be done through educational interventions e.g direct coaching. In terms of disease management to diabetic patients, it is addressed based on diabetes self-management (healthy diet, physical activity, monitoring blood sugar levels, medication adherence, problem solving, reduction of the complications risks, and effective coping (AADE, 2016) in Babalola, *et al*. 2021. It was further explained that nurse-led coaching in terms of lifestyle modification can reduce systolic blood pressure and reduce the risk of cardiovascular disease in the patients with type 2. In addition, the purpose of the health education is to increase the patient’s proactive involvement in self-care and increase adherence to medication and lifestyle changes (Lumu *et al*., 2021). The results of another study conducted by Baptista et al., 2019 showed that education conducted through training, face-to-face meeting with patients and home visits could increase knowledge in the management post prandial blood glucose levels, blood glucose peripherals, regulation of diet and drugs as well as menagament of drug administration for the prevention of hypoglycemia. Home visits aim to evaluate the patient’s activeness and involvement in the intervention provided so as to achieve the expected goals.

### Indirect methods of health education and its impact on type 2 diabetic patients

Indirect health education can be done through the use of mobile phones. In this case, nurse can develop smartphone-based education methods as a form of innovation to improve knowledge, attitudes and skills of type 2 diabetic patients in diabetes management. Asante *et a*l., 2020, further explained that health education interventions using mobile phones can increase the compliance of type 2 diabetic patients in self-care (diet, activity/exercise and foot care) and control HbA1c levels or improve short-to midle term glycemic management of diabetic patients with type2. This finding is supported by research conducted by (Dincer and Bahçecik, 2021) that health education using mobile phones has an impact on increasing knowledge, self-efficacy and lower limb/foot care behavior.

In providing indirect health education, nurse absolutely will take notice of various factors in order to support the successful implementation of health education. There are several factors that influence the success of implementing mobile phone-based health education, namely: resources (patient’s knowledge of using the application), perceptions of the severity of the disease and so on. These factors can be overcomed by providing a sort of training on the use of the application and adjusting perceptions with the patient on the method. In a study conducted by Asante *et al*., 2020, the application contained information about the patient’s self-care activities. Nurse in implementing smartphone-based education should make cell phone calls to patients to follow up on the patient’s self-care activities. Thus, all forms of the patient’s self-care activities are well monitored and this can improve the patients’ compliance in diabetes management. This is supported by research conducted by Jeffrey *et al*., 2019 which shows that the use of mobile phones can increase patient satisfaction due to interactions between patients and nurse. In addition, patients are also satisfied because they receive weekly messages related to self-care management sent via mobile phones.

### Mixed or combination methods of health education and its impact on type 2 diabetic patients

Mixed method of health education combined by nurses is one of the most beneficial interventions for type 2 diabetic patients. According to several research, health education provided directly by nurse and modified into smartphone-based health education will increase awareness of diabetic patients in performing self-care (Miyamato *et al*., 2019). Further explained by Alonso-Domínguez *et al*., 2019 that the combination of health education can increase knowledge in lifestyle modification (diet regulation, physical activities). In addition, there is a combination of educational methods such as training for patients and followed by monitoring using the telephone to evaluate the effectiveness of the methods used to increase knowledge about diabetes, improve self-care behavior and control HbA1c levels (Babalola *et al*., 2021). Through the use of smartphones, diabetic patients can contact or communicate with nurse regarding things they need and support in treatment. In addition, through health education provided by nurse, patients are given information about how to exercise simple physical activities that can reduce blood glucose levels and prevent various complications. Physical activity can be walking for 30 minutes and done 5 times per week. With the implementation of this health education, blood sugar levels are controlled and reduce the risk of diabetic complications.

## Conclusion

Health education is one form of learning that can be facilitated by nurses to increase knowledge, changing attitudes and behavior of patients in disease management, one of which is diabetes management. The health education can be exercised directly (training, coaching, individual teaching, FGD), indirectly (through the use of mobile phone applications) and a combination of both direct and indirect educational methods through mobile health technology and nurse health coaching or a combination of training and education. monitoring using phone calls, using smartphone applications and workshops on diet and activities management. All forms of health education exercised by nurse today are forms of nurse innovation to increase patients’ awareness, willingness and ability to increase self-care activities so adequate glycemic control can be achieved. Thus, various risks of complications due to diabetes can be minimized and, in the end, it will improve the quality of life in type 2 diabetic patients.

## Data Availability

All data produced in the present work are contained in the manuscript

